# Effect of HIV disease and the associated moderators on COVID-19 Mortality

**DOI:** 10.1101/2022.06.09.22275881

**Authors:** John Muthuka K., Francis Muchiri W., Kelly Oluoch J., Francis Muchiri W., Japheth Nzioki M.

## Abstract

**Introduction:** Established predictors for COVID 19 related mortalities are diverse. The impact of these several risk factors on coronavirus mortality have been previously reported in several meta-analyses limited by small sample sizes and premature data. The objective of this systematic review and meta-analysis coupled with meta-regression was to evaluate the updated evidence on the risk of COVID 19 related mortality by HIV serostatus using published data, and account for possible moderators.

**Method:** Electronic databases including Google Scholar, Cochrane Library, Web of Sciences (WOS), EMBASE, Medline/PubMed, COVID 19 Research Database, and Scopus, were systematically searched till 30th February, 2022. All human studies were included irrespective of publication date or region. Twenty-two studies with a total of 19,783,097 patients detailing COVID 19 related mortality were included. To pool the estimate, a random effects model with risk ratio as the effect measure was used. Moreover, publication bias and sensitivity analysis were evaluated followed by meta-regression. The trial was registered (CRD42021264761) on the PROSPERO register.

**Results:** The findings were consistent in stating the contribution of HIV infection for COVID-19 related mortality. The cumulative COVID-19 related mortality was 110270 (0.6%) and 48863 (2.4%) with total events of 2010 (3.6%), 108260 (0.5%) among HIV-positive and negative persons respectively. HIV infection showed an increased risk of COVID-19 related mortality [RR=1.19, 95% CI (1.02, 1.39) (P=0.00001)] with substantial heterogeneity (I squared > 80%). The true effects size in 95% of all the comparable populations fell between 0.64 to 2.22. Multiple Centre studies and COVID-19 mortality with HIV infection showed a significant association [RR = 1.305, 95% CI (1.092, 1.559) (P = 0.003)], similar to studies conducted in America (RR=1.422, 95% CI 1.233, 1.639) and South Africa (RR=202;1.123, 95% CI 1.052, 1.198). HIV infection showed a risk for ICU admission [(P=0.00001) (I squared = 0%)] and mechanical ventilation [(P=0.04) (I squared = 0%)] which are predictors of COVID-19 severity prior to death. Furthermore, risk of COVID 19 related mortality is influenced by the region of study (R squared = 0.60). The variance proportion explained by covariates was significant (I squared = 87.5%, Q = 168.02, df = 21, p = 0.0000) (R squared = 0.67).

**Conclusion:** Our updated meta-analysis indicated that HIV infection was significantly associated with an increased risk for both COVID 19 mortality, which might be modulated by the regions. We believe the updated data further will contribute to more substantiation of the findings reported by similar earlier studies (Dong et al., 2021; K. W. Lee et al., 2021; Massarvva, 2021; Mellor et al., 2021; Ssentongo et al., 2021)

## Introduction

By March 2022, there have been a cumulative 435,626,514 confirmed cases of COVID-19, including about 6 million deaths reported to WHO. SARS-CoV-2 infection may present asymptomatically or culminate to mild symptoms with a proportion of people developing severe coronavirus disease 2019 (COVID-19), which lead to hospitalization, acute respiratory distress syndrome (ARDS) or death. Established predictors for mortality are increasing age, male gender, dyspnea, hypertension and diabetes while severe COVID-19 is associated with hilar lymphadenopathy, bilateral lung and reticular pattern (Chidambaram et al., 2020).

About 38 million people globally living with HIV (PLWH), (including 1.7 million children), with a global HIV prevalence of 0.7% among adults (Vardell, 2020), may have an increased risk of adverse outcomes from COVID -19 infection as a result of HIV-associated immune dysfunction due to associated cells’ alterations and depletion (Korencak et al., 2019). There may also be a higher prevalence of co-morbidities among PLWH that predispose them to adverse COVID-19 outcomes (Mirzaei et al., 2021). Conversely, PLWH may have more favorable outcomes due to increased health awareness or close medical follow-up and constant reviews with some specific antiretroviral agents under consideration as potential treatments for COVID-19 (Pio Conti Ronconi G2, Caraffa A3, Gallenga CE4, Ross R5, 2020).

Severe COVID-19 disease manifested by fever and pneumonia, leading to acute respiratory distress syndrome (ARDS), has been described in up to 20% of COVID-19 cases. This is reminiscent of cytokine release syndrome (CRS)–induced ARDS and secondary hemophagocytic lymph histiocytosis (sHLH) observed in patients with SARS-CoV-2 (Moore & June, 2020), characteristics of CRS, including pulmonary inflammation, fever, and dysfunction of non-pulmonary organs. An increase in interleukin-6 in peripheral blood is a key risk factor and an early indicator of CRS in COVID-19. Both antibody and T cell responses are critical for effective control and clearance of SARS-CoV-2. More severe COVID-19 disease correlates with lymphopenia and low T cell concentrations(Chen et al., 2020; Lucas et al., 2020; Sekine et al., 2020).

COVID-19 associated mortality by HIV serostatus is not explicitly researched and most meta-analysis have focused on studies lacking comparator groups or they used a general population as controls unlike in the current study which restricts the comparator as HIV negative in the same included study. We aimed to evaluate the evidence regarding the risk of COVID-19 mortality in people with HIV (PWH) using both earlier and recently published data, and a meta-regression to ascertain the extent to which this risk is modified by other possible covariates.

## Methods

We utilized a systematic review to identify studies between 1^st^ April 2020 and 30^th^ February 2022 that described cytokine release syndrome and COVID -19 mortality in people with HIV (PLWH) and compared them with HIV-negative people, and, meta-analysis approach followed by a meta-regression, to ascertain the covariates associated with COVID-19 mortality. A standard search strategy was used in PubMed, and then modified according to each specific database to get the best relevant results. These included MEDLINE indexed journals; PubMed Central; NCBI Bookshelf, medRxiv, LitCovid, Trip, Google, Google Scholar and publishers’ Web sites. The basic search strategy was built based on the research question formulation (i.e., PICO or PICOS). They were constructed to include free-text terms (e.g., in the title and abstract) and any appropriate subject indexing (e.g., MeSH) expected to retrieve eligible studies, with the help of an expert in the review topic field or an information specialist. The summary of search terms was; COVID-19 severity OR mortality OR death; Corona Virus mortality OR death, etc. After some rounds of trial, refinement and formulation of the search term for PubMed as follows: (COVID-19 OR corona-virus virus OR coronavirus disease) AND (“the study” [Publication Type] OR “study as the topic” [MeSH Terms] OR “study” AND HIV serostatus AND [All Fields]). One author with extensive literature search experience and expertise performed the preliminary screening to exclude duplicates and studies not related to HIV infection. For remaining articles, another author performed title/topic and abstract screening, with subsequent full text review by two authors using a standardized data extraction form. Where disagreement was feasible, inclusion decisions were made by a third author. We also included preprints to capture the most recent and emerging evidence. Studies with 15 or less participants were excluded as they were less likely to have the power to detect meaningful relationships. The quality of the studies was evaluated using the Newcastle-Ottawa Scale for observational studies (Sidweli, 1993; Wells et al., 2000).

Observation studies reporting COVID-19-related death in people with and without HIV included in a meta-analysis. Specific relative risks (RRs) and hazard ratios were combined with random effects model to account for variability of the true effect between studies. To explore possible effect modifications, subgroup and meta-regression analyses were conducted for both COVID-19 related mortality. Meta-analysis was performed in Review Manager version 5.4 and Comprehensive Meta-Analysis (CMA) (version 3), (dichotomous data, random effects model), calculated the effect estimates as Risk Ratios (with 95% CI).

### Summary of included studies

We identified 1745 records and included a total of twenty-two detailing COVID-19 mortality as outcome in our final analysis (Fig. 1). The included studies were peer-reviewed with some, as preprints since the research quest sought to capture even the latest data and information.

**Fig. 1:**
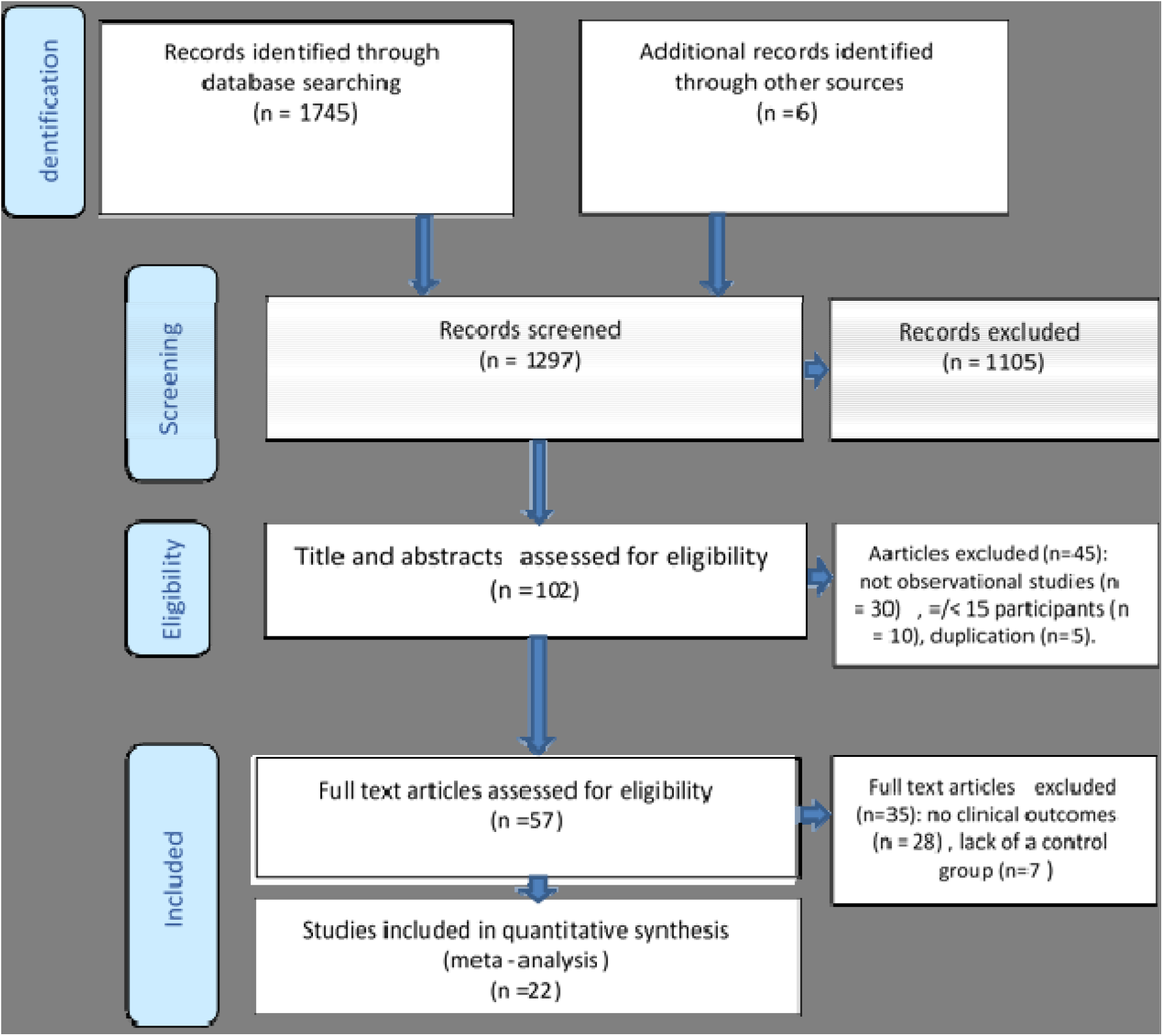
PRISMA flow diagram showing studies identified and included in a systematic meta-analysis of coronavirus disease 2019 Cytokine storm and mortality.

We identified twenty-one cohort studies (six prospective, fifteen retrospective) and, one case-control study comparing COVID-19-associated mortality between HIV Seropositive and HIV-sero-negative people, which we pooled in a meta-analysis (Bhaskaran et al., 2021; Braunstein et al., 2021; Chang et al., 2021; Díez et al., 2021; Durstenfeld et al., 2021; Geretti et al., 2021; Hadi et al., 2020; Karim et al., 2021; Karmen-Tuohy et al., 2020; Lee et al., 2022; Miyashita & Kuno, 2021; Nagarakanti et al., 2021; Patel et al., 2021; “Risk Factors for Coronavirus Disease 2019 (COVID-19) Death in a Population Cohort Study from the Western Cape Province, South Africa,” 2021; Sigel et al., 2020; Spinelli et al., 2021; Tesoriero et al., 2021; Venturas et al., 2021; Yang et al., 2021; Yendewa et al., 2021). Eleven of these (Braunstein et al., 2021; Chang et al., 2021; Díez et al., 2021; Durstenfeld et al., 2021; Hadi et al., 2020; Karmen-Tuohy et al., 2020; Lee et al., 2022; Nagarakanti et al., 2021; Patel et al., 2021; Yang et al., 2021; Yendewa et al., 2021) prior to mortality also, reported and compared cytokine release syndrome, defined by a specific parameter (such as intensive care unit admission) between HIV seropositive and seronegative.

In this meta-analysis pool, 19,783,097 from the 22 studies with mortality outcome (Bhaskaran et al., 2021; Braunstein et al., 2021; Chang et al., 2021; Díez et al., 2021; Durstenfeld et al., 2021; Geretti et al., 2021; Hadi et al., 2020; Harrison et al., 2020; Jassat et al., 2021; Karim et al., 2021; Karmen-Tuohy et al., 2020; Lee et al., 2022; Miyashita & Kuno, 2021; Nagarakanti et al., 2021; Patel et al., 2021; “Risk Factors for Coronavirus Disease 2019 (COVID-19) Death in a Population Cohort Study from the Western Cape Province, South Africa,” 2021; Sigel et al., 2020; Spinelli et al., 2021; Tesoriero et al., 2021; Venturas et al., 2021; Yang et al., 2021; Yendewa et al., 2021) were included. This inclusion utilized the predefined given CDC reporting guidelines on COVID-19 diagnosis (Chow et al., 2020). The cumulative COVID-19 mortality was 110270 (0.6%). The total COVID-19 mortality events were 2010 (3.6%) and 108260 (0.5%) the HIV sero-positive and HIV-seronegative persons respectively. The cumulative incidence of COVID-19 related mortality from all studies ranged from 0.09 % to 56 % (average: 14.4%). A summary of the studies included in this meta-analysis is available in Table 1.

**Table 1:** Summery of studies included in meta-analysis.

| First author | Location of patients | Study design and setting | Outcome (CRS or Mortality) | Events in HIV Seropositive/ total in Cohort | Events in HIV Sero-negative/total in cohort | Cumulative incidence of Mortality and CRS (defined by a specific parameter) (%) |
| --- | --- | --- | --- | --- | --- | --- |
| (Yang et al., 2021) | United States of America | Prospective Cohort, Cohort Collaborative (N3C) data Multiple sites | Mortality | 445 / 13170 | 25685 / 1423452 | 1.81885 |
| (Bhaskaran et al., 2021) | United Kingdom | Retrospective Cohort, Multiple sites | Mortality | 25685 / 1423452 | 14857 / 17255425 | 0.086108 |
| (Spinelli et al., 2021) | United States of America | Matched-Case-Control, Single site | Mortality | 5 / 1000 | 2 / 1000 | 0.35 |
| (Chang et al., 2021) | United States of America | Retrospective Cohort, Single site | Mortality | 1 / 10 | 223 / 1973 | 11.29602 |
| (Patel et al., 2021) | United States of America | Retrospective Cohort, Single site | Mortality | 22 / 100 | 1104 / 4513 | 24.40928 |
| (Durstenfeld et al., 2021) | United States of America | Prospective Cohort, Multiple sites | Mortality | 36 / 220 | 3290 / 21308 | 15.44965 |
| (Venturas et al., 2021) | South Africa | A retrospective analysis of prospectively collected data Cohort, Single | Mortality | 16 / 106 | 64 / 276 | 20.94241 |
| (Yendewa et al., 2021) | United States of America | Retrospective Cohort, Multiple sites | Mortality | 49 / 1638 | 6798 / 295556 | 2.303882 |
| (Braunstein et al., 2021) | United States of America | Retrospective Cohort, Multiple sites | Mortality | 313 / 2410 | 16160 / 202012 | 8.05833 |
| (Hadi et al., 2020) | United States of America | Prospective a propensity-matched cohort of patients without HIV Cohort, Multiple sites | Mortality | 20 / 404 | 1585 / 48178 | 3.303693 |
| (Harrison et al., 2020) | United States of America | Retrospective Cohort, Multiple | Mortality | 17 / 209 | 1279 / 29956 | 4.29637 |
| (Jassat et al., 2021) | South Africa | Retrospective Cohort, Multiple sites | Mortality | 644 / 2433 | 6122 / 26351 | 23.50611 |
| (Tesoriero et al., 2021) | United States of America | Retrospective Cohort, Multiple sites | Mortality | 207 / 2781 | 14522 / 360738 | 4.051783 |
| (Geretti et al., 2021) | United Kingdom | Prospective Cohort, Multiple sites | Mortality | 30 / 81 | 14555 / 28460 | 51.10192 |
| ("Risk Factors for Coronavirus Disease 2019 (COVID-19) Death in a Population Cohort Study from the Western Cape Province, South Africa," 2021) | South Africa | Retrospective Cohort, Single site | Mortality | 115 / 3863 | 510 / 17820 | 2.882442 |
| (Díez et al., 2021) | Spain | Retrospective Cohort, Multiple sites | Mortality | 2 / 21 | 12 / 105 | 11.11111 |
| (Karmen-Tuohy et al., 2020) | United States of America | Retrospective Cohort, Multiple sites | Mortality | 6 / 21 | 10 / 42 | 25.39683 |
| (Karim et al., 2021) | South Africa | longitudinal observational cohort study | Mortality | 13/93 | 8 / 143 | 8.898305 |
| (Sigel et al., 2020) | United States of America | Prospective Cohort Study, Single | Mortality | 18/88 | 81/405 | 20.08114 |
| (Miyashita & Kuno, 2021) | United States of America | Retrospective Cohort, Multiple sites | Mortality | 23/161 | 1235/8751 | 24.40928 |
| (Lee et al., 2022) | United Kingdom | Prospective Cohort Study, Multiple sites | Mortality | 0/17 | 5/50 | 7.462687 |
| (Nagarakanti et al., 2021) | ISRAEL | Retrospective Cohort, Single Site | Mortality | 3/23 | 153/254 | 56.31769 |
| Cumulative COVID-19 Mortality, all studies (%) |  |  |  |  |  | 110270 / 19783097 (0.557395 %) |

### Quality of evidence and risk of bias assessment

Included in Table 2 is the Newcastle-Ottawa scale for quality assessment and risk of bias. We assessed the quality of the included studies based on a modified version of the Newcastle-Ottawa Scale (NoS) which consists of 8 items with 3 subscales, and the total maximum score of these 3 subsets is 9. We considered a study which scored ≥7 a high-quality since a standard criterion for what constitutes a high-quality study has not yet been universally established. The twenty-two studies (with eleven of them having both CRS and mortality measurements making a total of thirty-three) assessed by us generated a mean value of 6.59 and as a result, overall quality was found to be moderate (NOS score min: 5, max: 8).

**Table 2:**
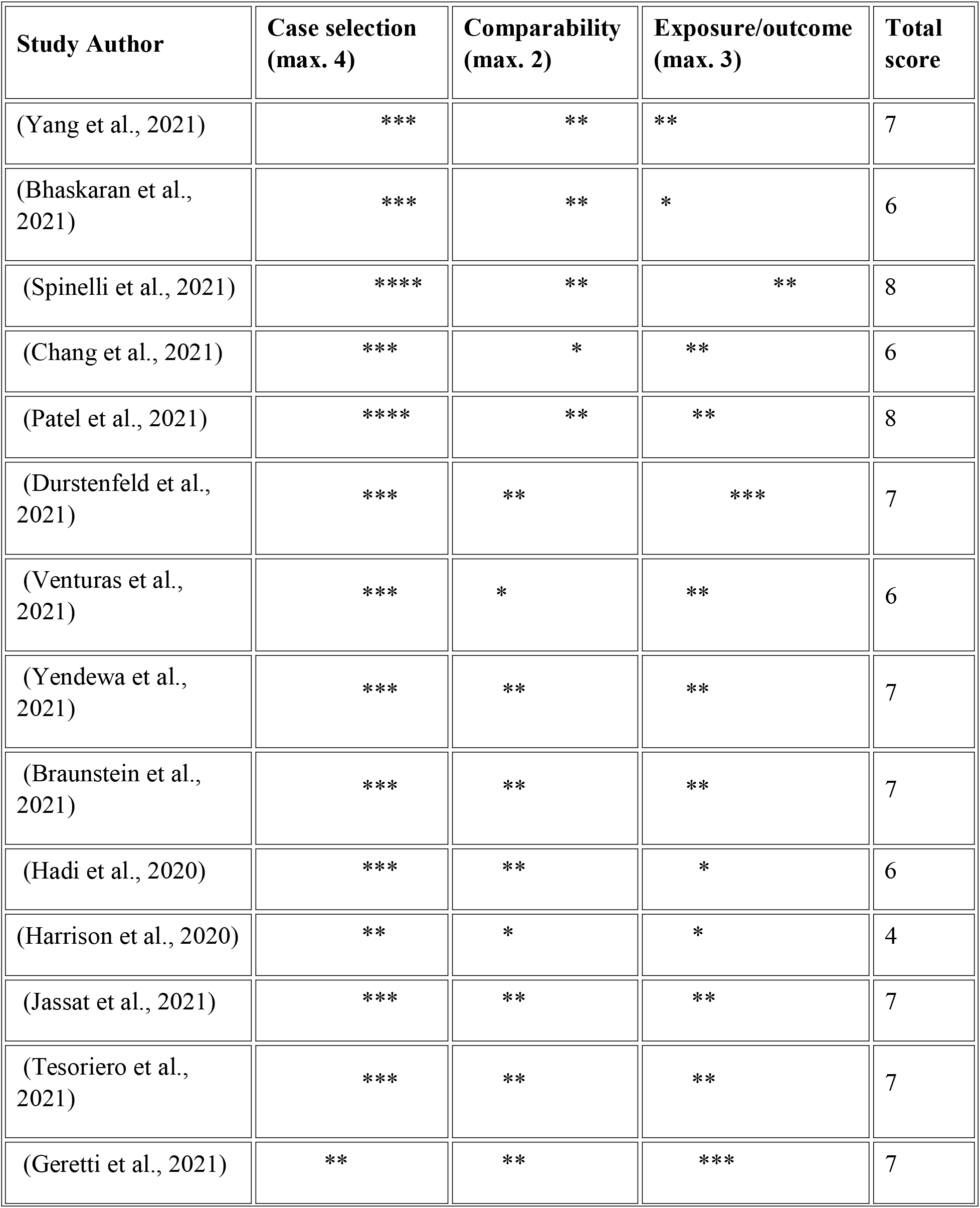
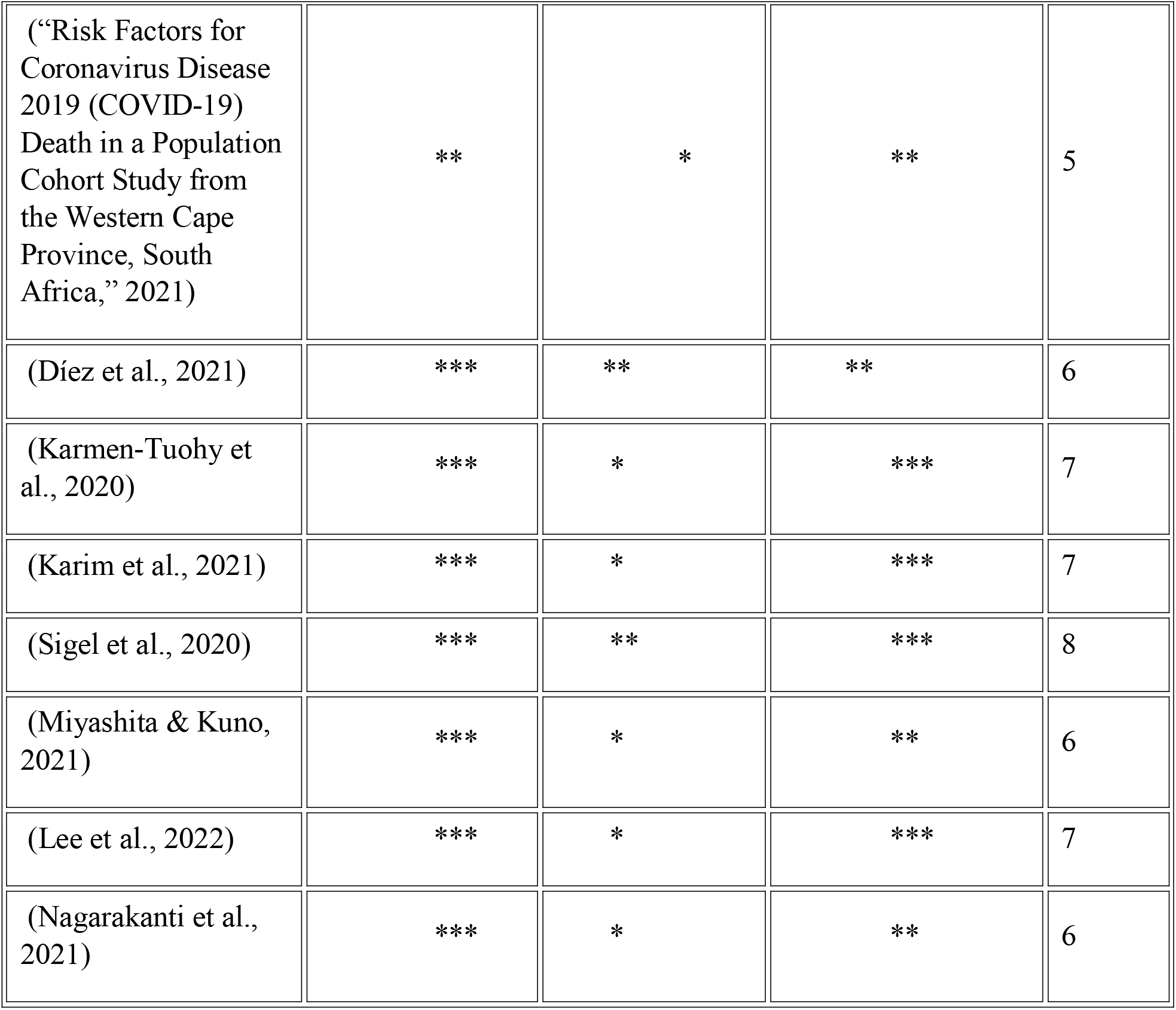
Newcastle-Ottawa scale for quality assessment and risk of bias.

There were common limitations among the included studies. Most were retrospective analyses of routinely collected clinical data, meaning identification of COVID-19 cases was not systematic and depended on the local approach to screening and diagnosis with only five prospective cohort studies. This may have varied over time and between settings, and may also differ between PLWH and the general population but in the case of this study though, the studies included both HIV-seropositive and seronegative populations’ data. Across all studies, the numbers of HIV-seropositive and COVID-19 infection were relatively low.

### Cumulative analyses of HIV-serostatus and the risk of COVID-19 related mortality

In this meta-analysis of the twenty-two studies (Bhaskaran et al., 2021; Braunstein et al., 2021; Chang et al., 2021; Díez et al., 2021; Durstenfeld et al., 2021; Geretti et al., 2021; Hadi et al., 2020; Harrison et al., 2020; Jassat et al., 2021; Karim et al., 2021; Karmen-Tuohy et al., 2020; Lee et al., 2022; Miyashita & Kuno, 2021; Nagarakanti et al., 2021; Patel et al., 2021; “Risk Factors for Coronavirus Disease 2019 (COVID-19) Death in a Population Cohort Study from the Western Cape Province, South Africa,” 2021; Sigel et al., 2020; Spinelli et al., 2021; Tesoriero et al., 2021; Venturas et al., 2021; Yang et al., 2021; Yendewa et al., 2021) that accounted for COVID-19 mortality as the outcome, the risk for HIV-seropositive persons was at an excess of about 20% with a significant difference between the studies [Heterogeneity: Tau^2^ = 0.08; Chi^2^ = 168.59, df = 21 (P < 0.00001); I^2^ = 88%] [Risk ratio = 1.19, 95% confidence interval (CI) 1.02 - 1.39] (Fig. 2). The prediction interval (Fig 3) demonstrates the true effects size in 95% of all the comparable populations falling between 0.64 to 2.22 demonstrating that, in some populations, the risk of COVID-19 mortality due to HIV infection is at one extreme of effect as low as 0.64 and as high as 2.22. This diversity informed the need to account for possible moderators. Thirteen of these studies were conducted in the united states of America (Braunstein et al., 2021; Chang et al., 2021; Durstenfeld et al., 2021; Hadi et al., 2020; Harrison et al., 2020; Karmen-Tuohy et al., 2020; Miyashita & Kuno, 2021; Patel et al., 2021; Sigel et al., 2020; Spinelli et al., 2021; Tesoriero et al., 2021; Yang et al., 2021; Yendewa et al., 2021)and the rest from other parts of the globe especially the United Kingdom and South Africa. A precision funnel plot for publication bias revealed considerable true heterogeneity between all the pooled studies (I^2^ = 88 %; P < 0.00001) (Fig 4). The egger’s regression test indicated no publication bias (intercept = - 1.10498, 95% confidence interval (−2.85566, 0.64570), with t=1.31660, df=20 and 1-tailed P = 0.10143). A sensitivity analysis was performed to explore the impact of excluding or including studies in meta-analysis based on sample size, methodological quality and variance on the heterogeneity obtained (I^2^ = 88 %). Elimination of six studies (Braunstein et al., 2021; Geretti et al., 2021; Harrison et al., 2020; Nagarakanti et al., 2021; Tesoriero et al., 2021; Yang et al., 2021) which were the major contributors of heterogeneity significantly reduced it [ Heterogeneity: Tau^2^ = 0.00; Chi^2^ = 15.90, df = 15 (P = 0.39); I^2^ = 6% ] (Fig 5). A funnel plot of precision (Fig 6), with egger’s test (intercept (= -0.16647, 95% confidence interval (−0.92034, 0.58740), with t=0.47361, df=14 and 1-tailed P = 0.32154) further indicating no publication bias.

**Figure 2:**
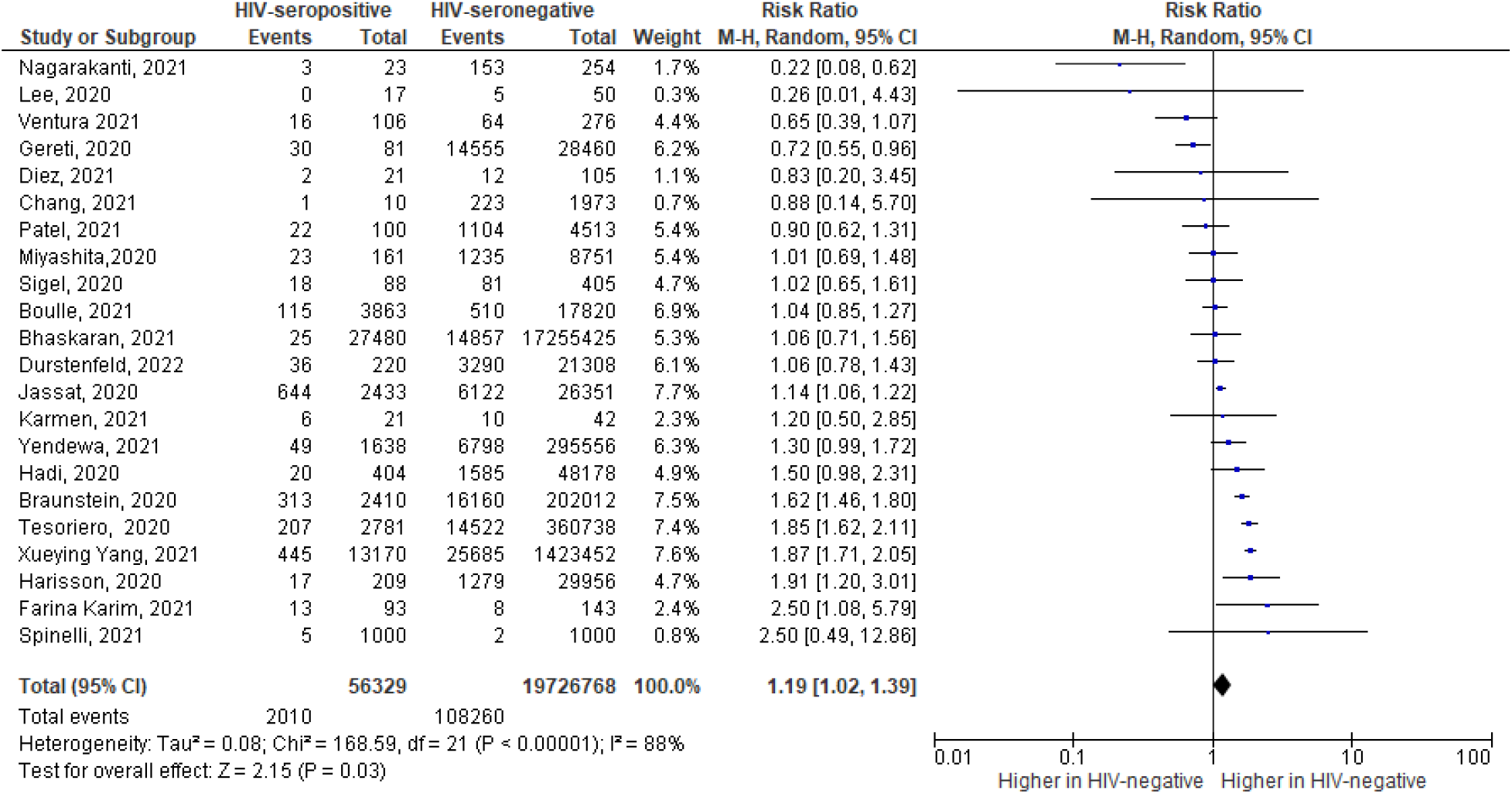
A forest plot of HIV-serostatus and the risk of COVID-19 related mortality.

**Figure 3:**
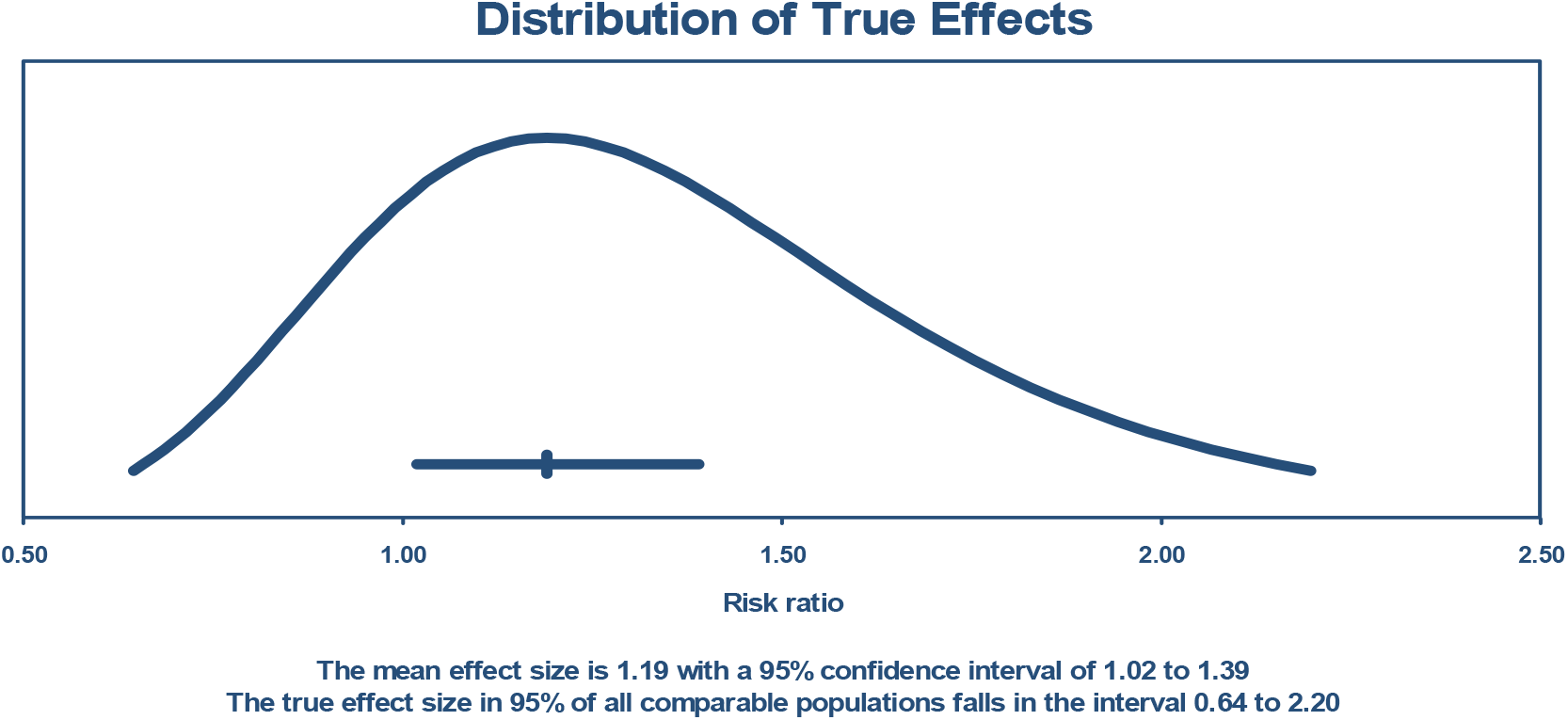
The prediction interval for the true effect size in 95% comparable population.

**Figure 4:**
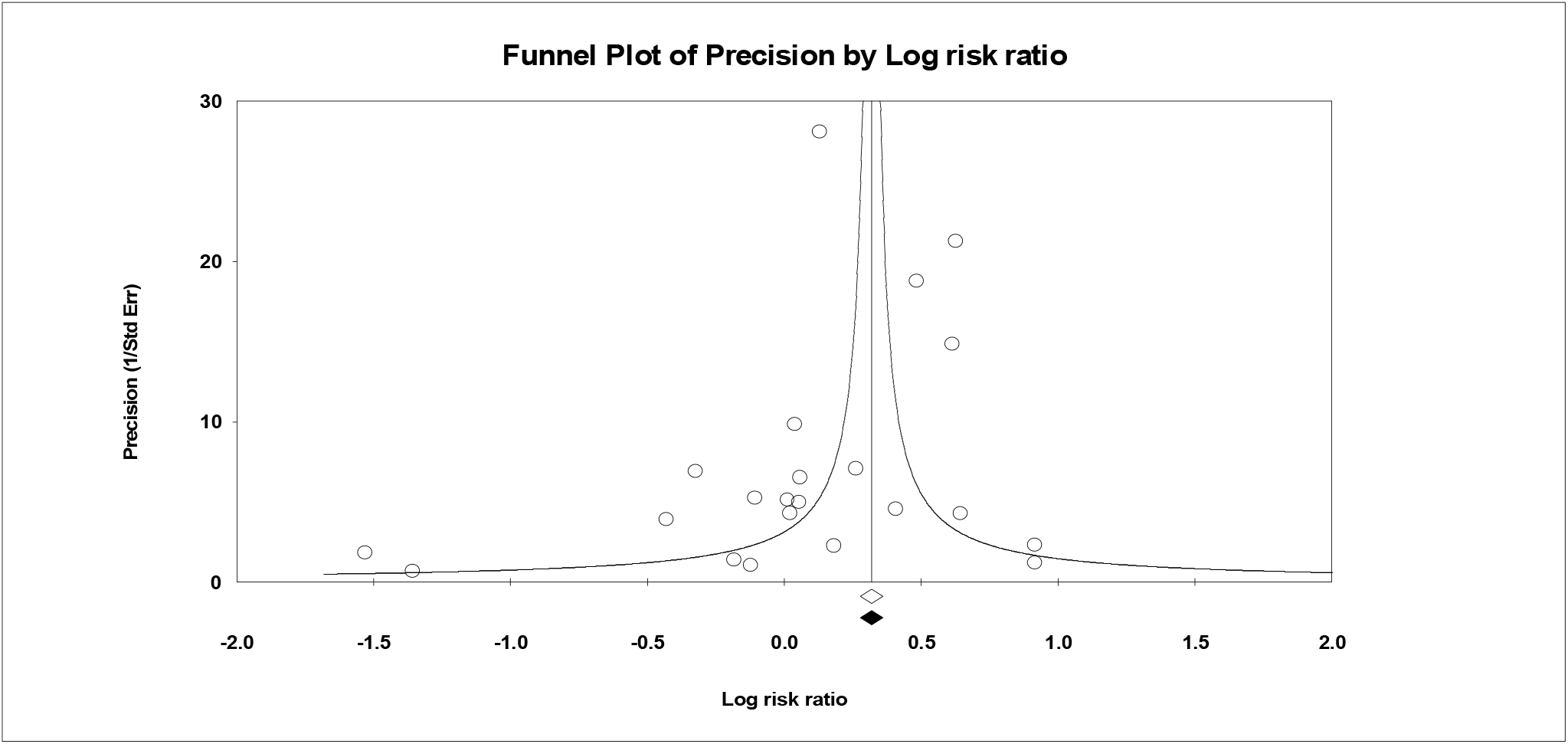
A precision funnel plot for publication bias.

**Figure 5:**
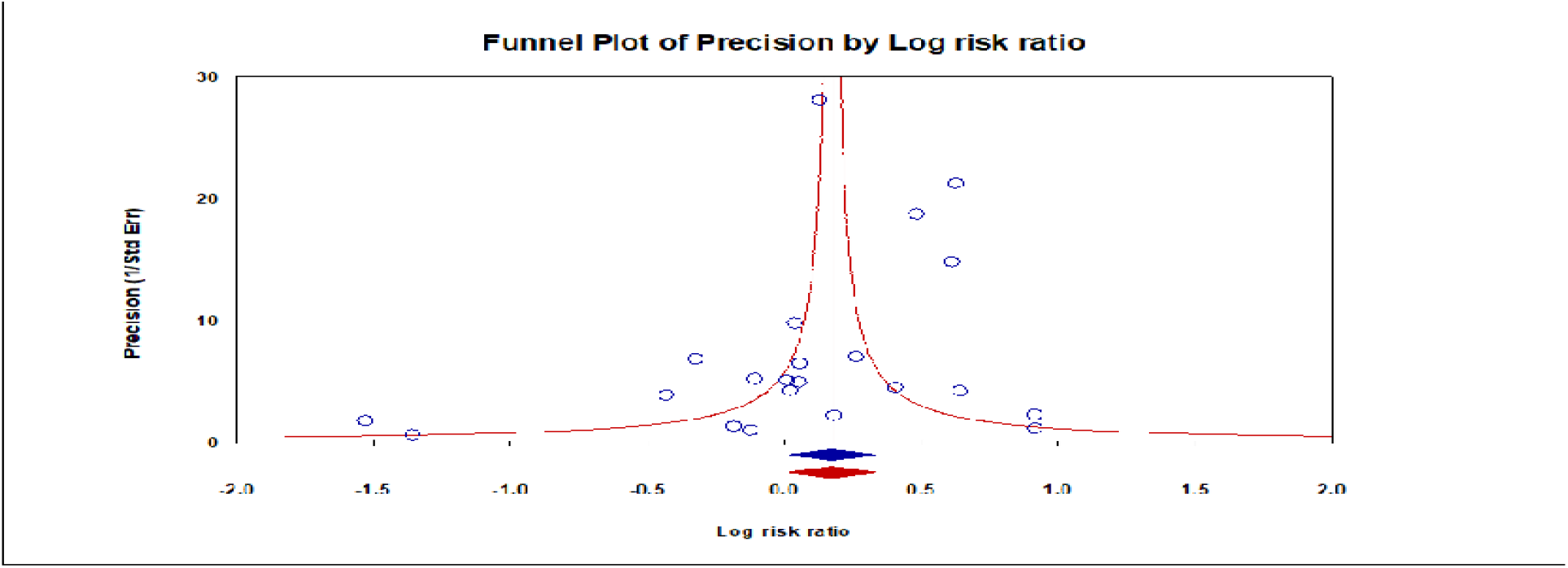
A sensitivity analysis to account for the heterogeneity.

**Figure 6:**
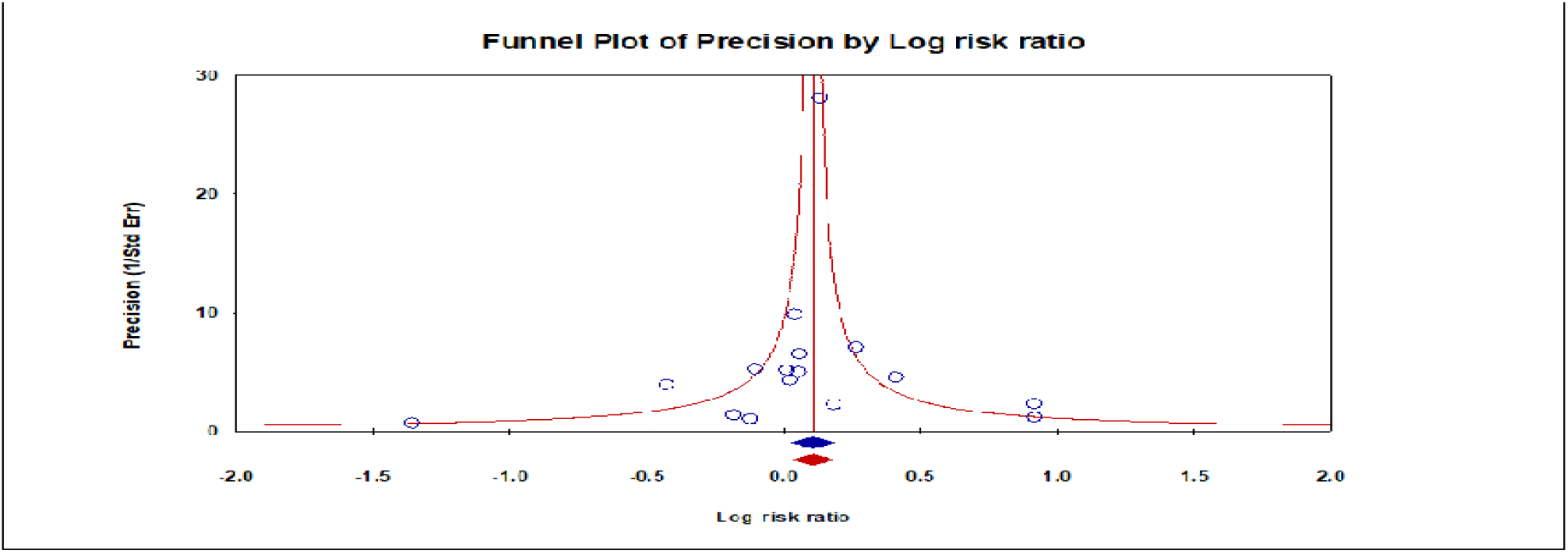
A funnel plot of precision with egger’s test for publication bias.

### Heterogeneity Investigation on HIV-serostatus and the risk of COVID-19 related mortality

We conducted a subgroup analysis on the nature of a study setting or number of sites which showed that, multiple Centre studies had a higher risk for COVID-19 mortality with HIV seropositive status [Risk ratio = 1.31, 95% confidence interval (CI) 1.09 -1.56] (P = 0.003), compared to single Centre studies (P = 0.67) (Fig 7). Independent sensitivity analysis clearly revealed, single Centre studies majorly contributed to the heterogeneity after the only four studies conducted in multiple sites (Braunstein et al., 2021; Geretti et al., 2021; Tesoriero et al., 2021; Yang et al., 2021) were removed from the analysis with non in single Centre studies reducing total heterogeneity (I^2^_=_ 40.5%) Heterogeneity: Tau^2^ = 0.00; Chi^2^ = 9.13, df = 9 (P = 0.43); I^2^ = 1% and Heterogeneity: Tau^2^ = 0.09; Chi^2^ = 17.09, df = 7 (P = 0.02); I^2^ = 59% respectively (Fig 9).Analysis by study design grouping did not show any statistical significance with HIV seropositive status with all the three clusters (prospective, retrospective cohorts and case control) (P < 0.05) (Fig 10). By region, HIV infection had a higher risk of COVID-19 mortality than those without HIV infection in America (RR□=□1.422, 95% CI 1.233–1.639), South Africa from 4 studies (RR□=□1.123, 95% CI 1.052–1.198) but not in the United Kingdom (RR□=□0.819, 95% CI 0.651–1.1030), Spain and Israel (Fig 7).

**Figure 7:**
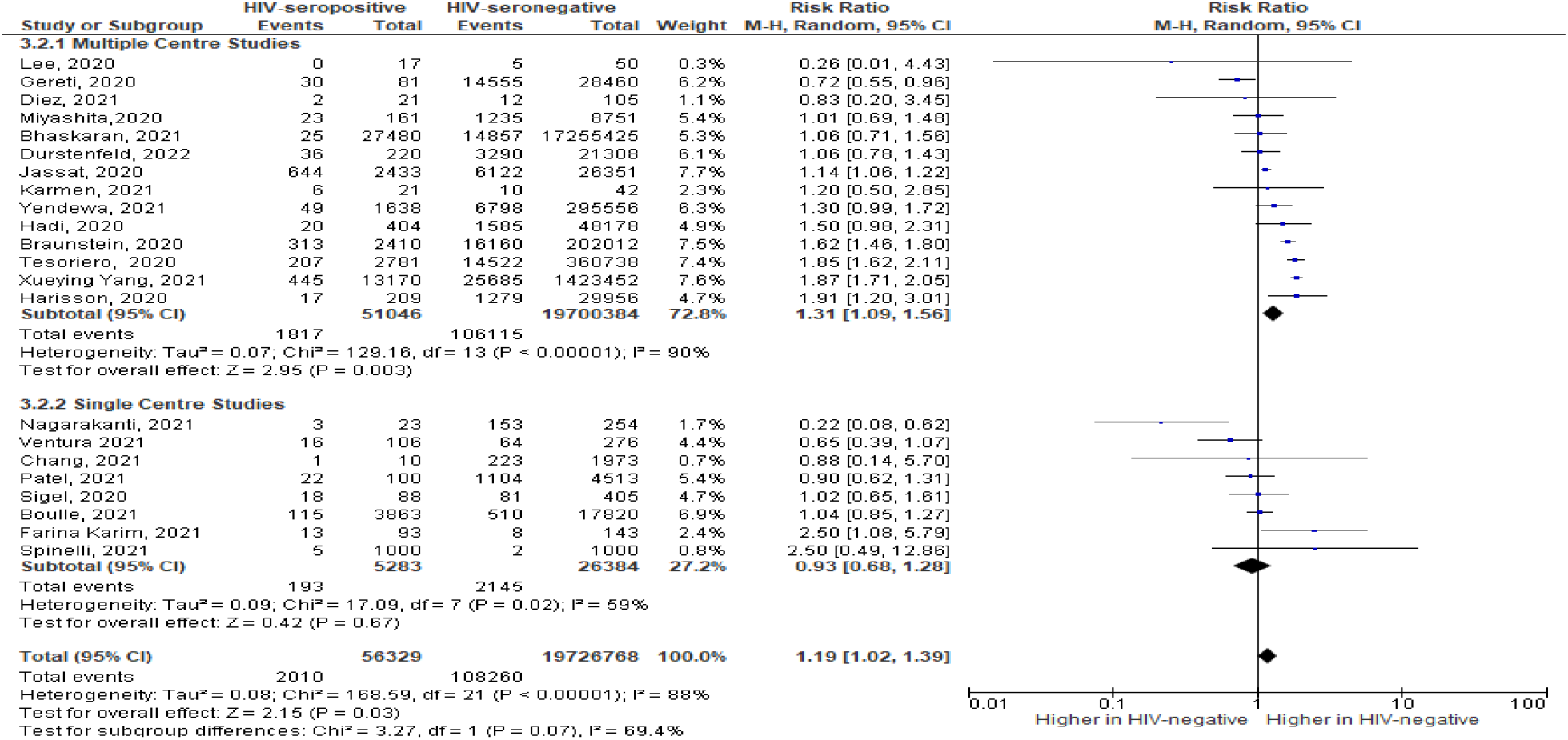
Subgroup analysis on the nature of a study setting or number of sites for heterogeneity investigation.

**Figure 8:**
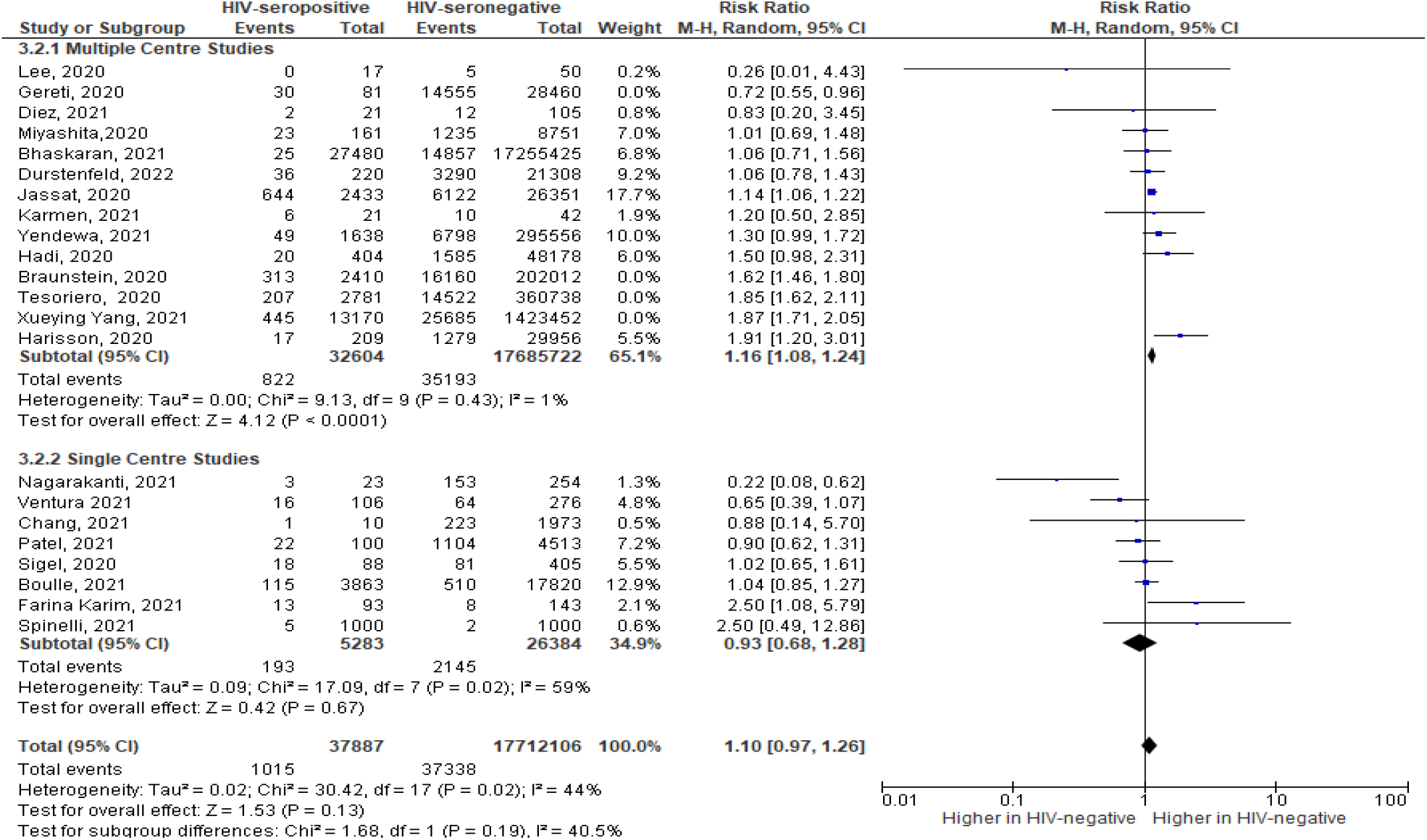
Independent sensitivity analysis on study setting.

**Figure 9:**
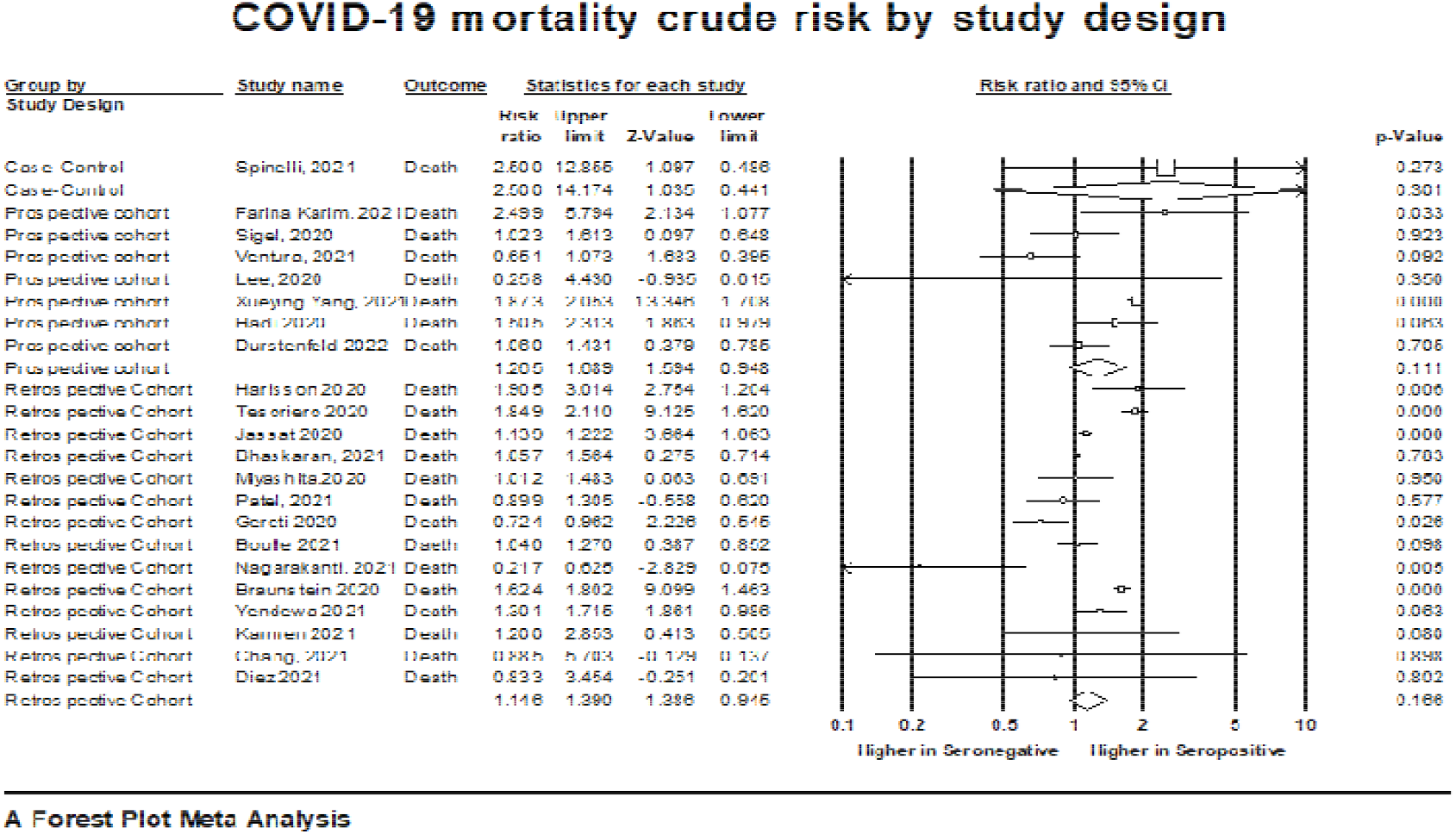
Analysis by study design grouping for heterogeneity investigation.

**Figure 10:**
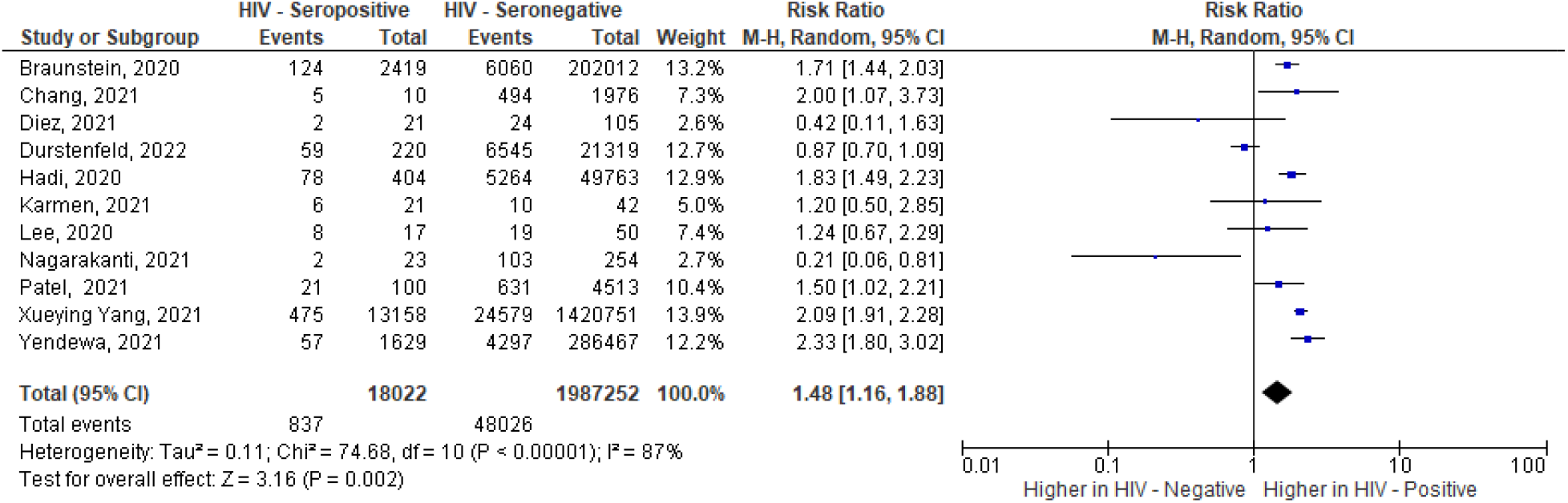
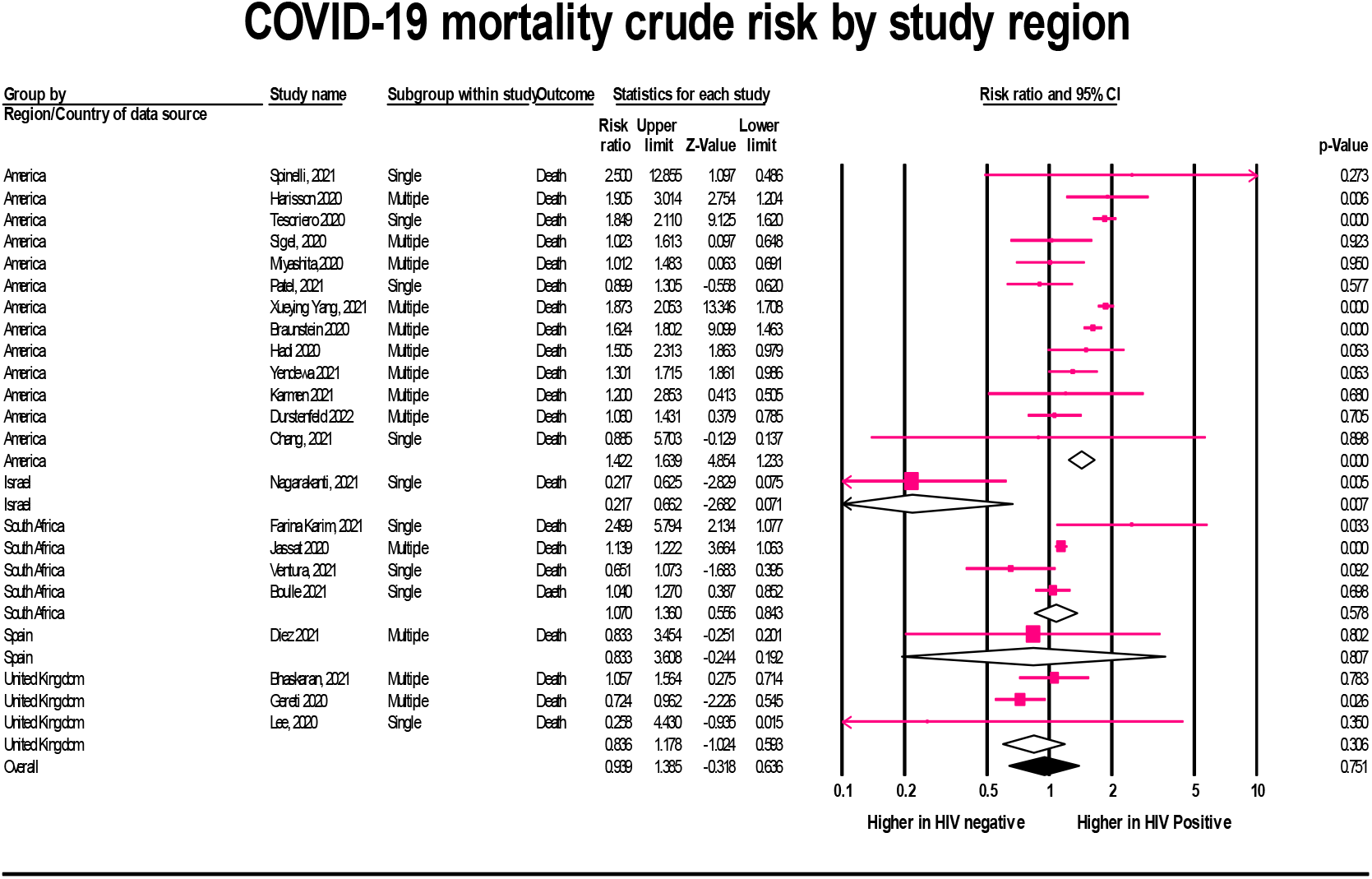
Heterogeneity Investigation by Study region.

### Meta-regression for possible moderators of coronavirus disease mortality with HIV-serostatus

The value of I^2^ in studies accounting for COVID-19 related mortality was 88, which means that, over 80% of the observed variance came from real differences between studies and, as such, can potentially be explained by study-level covariates. To ascertain this, the analysis assessed the possible influence of pre-determined moderators.

Moderation analysis for studies detailing COVID-19 mortality revealed no association with the nature of study site (single or multiple) (Q = 3.58, df = 1, p = 0.0584) (R^2^ = 0.09) (Supp. File 1). The region / country of study population was associated with COVID-19 mortality (Q = 19.71, df = 4, P = 0.0006) (R^2^ = 0.60) (Supp. File 2)., while the year of the study did not independently predict COVID-19 mortality (Q = 0.70, df = 2, P = 0.7043) (Supp. File 3). The combined impact of all covariates in the model explained at least some of the variance in effect size (Q = 28.84, df = 7, P = 0.0002). Further, the proportion of variance explained by covariates on comparing the model with and without the covariates was significant (Tau^2^ = 0.0834, Tau = 0.2887, I^2^ = 87.5%, Q = 168.02, df = 21, p = 0.0000) (R^2^ = 0.67) (Supp. File 4).

## Discussion

The purpose of this study was to systematically review and conduct meta-analysis using the most current data from studies on the incidence of COVID-19 related mortality relative to HIV serostatus, alongside the associated covariates via meta-regression. Further, it aimed at ascertaining the combined proportion effect of all covariate in studies detailing mortality.

To our knowledge, this is one unique meta-analysis amongst peer-reviewed literature assessing the risk of HIV on the incidence COVID -19 related mortality by further investigating the associated moderators of the same. Principally, the present meta - analysis found that HIV seropositive status portrays an increased risk for COVID-19 related mortality at approximately twenty percent in comparison to HIV seronegative counterpart population. Overall, there was a high degree of heterogeneity amongst studies detailing the COVID-19 related mortality, which greatly reduced following sensitivity analysis. The outcome remained significant on inclusion of only good-quality studies suggesting these analyses represent true effects as per the generated prediction intervals. A high level of heterogeneity was only observed with inclusion of few studies in assessing the effect of HIV on COVID-19 related mortality, likely to substantial inter-study variation. The egger’s regression test indicated low impact of publication bias on our results.

The cumulative incidence of mortality with HIV seropositive status was at an average excess of 20% (RR=1.197), this was similar to a recent meta-analysis which reported almost same results in odds ratio suggesting that, people living with HIV infection had a higher risk of mortality from COVID-19 than those without HIV infection (OR□=□1.252, 95% CI 1.027–1.524)(Dong et al., 2021). Among the COVID-19 patients with HIV infection, the mortality rate due to COVID-19 was 3.6 %, and among the COVID-19 patients without HIV infection, the mortality rate due to COVID-19 was 0.5% this is commensurate with a meta-analysis findings that showed a 3.44% and 0.42% respectively(Dong et al., 2021).

Our finding that HIV infection is associated with increased COVID-19 related mortality further validates previous research findings from several smaller meta-analyses and primary studies (Davies, 2020; Dong et al., 2021; Mellor et al., 2021; Ssentongo et al., 2021) in a most recent data from a larger patient population, achieved through a more rigorous, prospectively registered methodology. The association of the COVID-19 mortality with HIV seropositive status in the context of this current findings is biologically plausible as in normal circumstances, as ARDS leads to COVID-severity prior to death (Hu et al., 2021).

Alongside other co-morbidities, HIV has been found to advance the clinical severity of COVID-19 culminating to death (Essien et al., 2021). In addition, HIV infection increases severity from both bacterial and viral infections through the induction of mechanical and structural changes in the respiratory tract and alteration of cell and humoral-mediated immune responses(Eybpoosh et al., 2021). In the context of respiratory viruses, HIV infection has been reported to cause increased hospital and ICU admissions with influenza infection, greater severity with respiratory syncytial virus bronchiolitis and increased mortality with viral pneumonia (Garbino et al., 2008).

Analysis of HIV infection to the risk of COVID-19 mortality by the study setting showed significance with multiple Centre studies [RR = 1.305, 95% CI 1.092 -1.559] (P = 0.003) an outcome similar to a multicenter meta-analysis (Dandachi et al., 2021; Feng et al., 2020; Liu et al., 2020). With hypothesis that studies in our meta-analysis stemmed from one similar region with its own overall true effect on sub-subgroup analysis, fixed effect model was used and people with HIV infection had a higher risk of COVID-19 mortality than those without HIV infection in the United States (RR□=□1.422, 95% CI 1.233–1.639). A similar risk was also achieved in South Africa from 4 studies, our subgroup analysis found that people living with HIV infection also had a higher risk of COVID-19 mortality than those without HIV infection (RR□=□1.123, 95% CI 1.052–1.198). However, no significant association between HIV infection and the mortality risk of COVID-19 was found in the United Kingdom (RR□=□0.819, 95% CI 0.651–1.1030). This finding is very consistent with another meta-analysis which showed; USA (OR□=□1.520, 95% CI 1.252–1.845), South Africa (OR□=□1.122, 95% CI 1.032–1.220) and United Kingdom (OR□=□0.878, 95% CI 0.657–1.174). The reason for this difference might be related to SARS-CoV-2 virus mutations in different countries (Nagy et al., 2021; WHO, 2021). Further, access and affordability of health services in different countries could also affect(Amimo et al., 2021; Singh et al., 2021).

Meta-regression analysis showed that, the year (2020, 2021 and not 2022) and the region a study was conducted were associated with COVID-19 related mortality (P<0.05), unlike the study setting sites. This trend has been explained by the data on cumulative mortality globally where the case fatality rates were higher in 2020 and in a reducing trajectory in 2021 and a flattening curve in 2022 (Horwitz et al., 2021; James & Menzies, 2021). Generally, the combined impact of all covariates in the model explained at least some of the variance in COVID-19 related mortality, similar to existing findings in countries and region related factors (Asfahan et al., 2020; Hashim et al., 2020). Further studies show that, patients located in America experienced higher mortality rates from COVID-19 infection (aOR, 7.441; 95% CI, 3.546-15.617) (Albitar et al., 2020), which in the context of this study, formed the largest population from 13 of all the twenty two studies.

## Conclusion

Our study indicated a consistent and statistically significant effect of HIV on COVID-19 related mortality even after heterogeneity investigation all in random effects model with egger’s regression test indicating no major publication bias. With a fixed effect model, region of study greatly influenced COVID-19 related mortality alongside HIV infection. The proportion of variance explained by covariates was significant with the year and region of study being the major co-variates associated with both COVID-19 related mortality. Public health interventions should be carefully tailored and implemented on HIV infected persons with COVID□19 to reduce the risk of severity associated mortality. An intensive and regular focus is required to detect early occurrences of clinical conditions in similar viral pandemics or COVID-19 resurgence.

## Data Availability

All data produced in the present study are available upon reasonable request to the authors

## Acknowledgements

Author contributions: **J.M.K**. conceived the study. **MJ**. performed the literature search. **J.M.K**. and **F.M.W**. contributed equally to the screening and qualitative analysis, with support from J.K.O. and **M.J. J.M.K**. performed the meta-analysis. **J.M.K**. and **J.K.O**. wrote the first draft of the article. **J.M.K**., **J.K.O**., **MJ**., **F.M.W**., critically reviewed and edited the article and consented to publication.

